# Ultrasonography in General Practice: insights from an implementation study in a Low-income neighborhood of Amsterdam

**DOI:** 10.1101/2025.05.22.25327578

**Authors:** Ralf E. Harskamp, Britt Soddemann genaamd Keute, Constantijn Pleiter, Jelle C.L. Himmelreich

**Author notes:** Corresponding author Name: Dr. Ralf Harskamp, E-mail address, Department: General Practice, Institute: Amsterdam UMC. authors share first authorship due to equal contribution to the manuscript (Ralf E. Harskamp, Britt Soddemann genaamd Keute).

## Abstract

**Background:** Ultrasonography is increasingly being used in primary care settings, facilitated by technological advancements, reduced device-related costs, and improved access to training and support. It holds particular promise for underserved communities, where barriers such as co-pays or long waiting lists can limit access to diagnostic services. This study evaluates the impact of a primary care ultrasonography service in an underserved urban community.

**Methods:** We conducted a retrospective cohort study of consecutive patients who underwent ultrasonography between 2019-2024 at a large primary care center (n=7877 patients) in Amsterdam the Netherlands. The primary outcomes included the frequency, indications, and diagnostic outcomes. Secondary outcomes included diagnostic accuracy, changes in clinical decision-making, referral rates, and costs.

**Results:** A total of 394 patients were included, of whom 62.4% underwent abdominal ultrasonography, 31.7% gynecological ultrasonography, and 5.8% other types of ultrasounds. Ultrasonography yielded conclusive results in 83.5% of cases, and reduced secondary care referrals by 58.8%. The sensitivity and specificity for ultrasonographic abnormalities were 98.9% (95%CI: 94.2-99.9%) and 89.8% (95%CI: 85.3-93.4%), respectively.

**Conclusion:** Primary care ultrasonography enhances diagnostic efficiency and reduces unnecessary referrals in more than half of cases. Its use in underserved communities can improve healthcare access and help address system-related health disparities. Further research is needed to assess its cost-effectiveness and scalability.

## Introduction

### Background

Ultrasonography is a widely used, non-invasive diagnostic tool employed across various medical disciplines. Traditionally confined to hospitals and specialized diagnostic centers, ultrasonography is increasingly utilized in primary care.(1, 2) This expansion is driven by technological advances, such as portable devices, artificial intelligence support, and high-quality training programs.(1-3) The availability of primary care ultrasonography could lower the threshold to ultrasound imaging, particularly in underserved communities, e.g. by lowering waiting time and by reducing co-pay in setting where this is relevant. In the Netherlands, where such co-pays apply to secondary care imaging, healthcare insurers reimburse general practitioners (GPs) for performing ultrasonograms and do not charge a co-pay to patients, which makes a primary care ultrasonography service of particular relevance in underserved communities.(3-5) Despite the growing uptake, limited attention has been paid to the use and impact of ultrasonography in primary care, certainly not in low-income communities. In this study we therefore set out to study these aspects, in which we evaluated indications, diagnostic outcomes, and impact on treatment and referral patterns in a large primary care health center in a socioeconomically disadvantaged community in Amsterdam, the Netherlands.

### Methods

For this study we reported our findings according to ‘standards for reporting implementation studies, StaRI’.(6)

### Study design & setting

We conducted a retrospective cohort study of consecutive patients who underwent ultrasonography at Gezondheidscentrum Holendrecht, a large primary care health center in a socio-economically disadvantaged area in Amsterdam, the Netherlands. The health center serves a population of almost eight thousand patients, a large proportion of whom have a non-western migration background. A total of 15 salaried GPs worked at the health center during the study period, four of whom were certified to perform ultrasonograms. We collected data through electronic health record (EHR) review between May and July of 2024 and involved data from patients who underwent ultrasonography between January 2019 and June 2024.

### Participants

The study included all patients who underwent primary care ultrasonography as indicated by their treating GP, according to following the health center’s inclusion criteria which included primarily abdominal or gynecological indications. We applied no further exclusion criteria for inclusion in the current analysis beyond patients who opted out of scientific research participation.

### Ultrasonography

Ultrasonography procedures were integrated into the primary care practice following the implementation of a structured protocol. Training for GPs included completion of a certified ultrasonography course and subsequent registration in a national register for primary care echographers, which also requires regular practice, use of a structured scanning protocol, and training to ensure recertification. The GPs documented each primary care ultrasonogram using standardized digital forms within the EHR.

### Data collection

From the EHR we extracted data on patient demographics, medical history, clinical symptoms, primary care ultrasonogram indications and results, number of referrals to secondary care, results of secondary care imaging including ultrasonography, as well as data on clinical outcomes during follow-up.

### Outcome measures

Outcomes reported in this analysis involved the frequency, indications and subsequent clinical course of primary care ultrasonograms. Diagnostic accuracy for overall ultrasonogram, as well as for the most common conditions, gallstones and kidney stones, were evaluated. As the reference for diagnostic accuracy analyses we used a secondary care ultrasonogram or, in cases where no secondary care ultrasonogram was performed, we manually reviewed the patient’s EHR for clinical follow-up related to the index primary care ultrasonogram.

### Statistical analysis

We reported descriptives of categorical data as numbers and percentages, and of continuous data as median and interquartile range (IQR). We compared proportions using the Fisher exact test or Pearson χ^2^ test and continuous variables using Student t test in case of normally distributed data or Mann–Whitney U test in case of non-normally distributed data. We assessed normality of distribution of continuous data using the Q-Q plot and Kolmogorov–Smirnov test. We evaluated statistical significance in all analyses at the .05 level using two-tailed tests. We used SPSS for descriptive analyses of patient characteristics and clinical outcomes.(7) In our diagnostic accuracy analyses we provided sensitivity, specificity, positive and negative predicting values (PPV & NPV, respectively) and positive and negative likelihood ratio (LR+ and LR-, respectively) with 95% confidence interval (95%CI) using MedCalc version 23.0.6.(8) A test with an LR+ over 10 or LR-under 0.1 is generally considered to be reliable for diagnosing and ruling out a condition, respectively.(9)

### Ethics

We captured data in an anonymized electronic case report form. The use of deidentified routine primary care data for research purposes was granted a waiver for informed consent (W23_074).

### Patient and public involvement

Patients or the public were not involved in the design, or conduct, or reporting, or dissemination plans of our research.

## Results

### Participant Characteristics

The study population included 394 patients, of whom 70.6% were female. The median age for male patients was 57 years (IQR: 39-70), and for female patients, 44 years (IQR: 29-61). Cardiovascular disease risk factors were common in the cohort, particularly among patients undergoing abdominal ultrasonograms, as displayed in *table 1*.

**Table 1.** Patient characteristics of the total cohort as well as stratified by indication.

|  | *Total* | *Abdominal*<br>*ultrasounds* | *Gynecological*<br>*ultrasounds* | *Other*<br>*ultrasounds* |
| --- | --- | --- | --- | --- |
| *Number of patients* | 394 (100%) | 246 (100%) | 125 (100%) | 23 (100%) |
| *Relevant medical history* |  |  |  |  |
| *None* | 160 (40.6%) | 83 (33.7%) | 66 (52.8%) | 11 (47.8%) |
| *Hypertension* | 83 (21.1%) | 62 (25.2%) | 17 (13.6%) | 4 (17.4%) |
| *Hypercholestrolemia* | 34 (8.6%) | 23 (9.3%) | 8 (6.4%) | 3 (13.0%) |
| *Cardiovascular diseases\** | 27 (6.9%) | 23 (9.3%) | 2 (1.6%) | 2 (8.7%) |
| *Diabetes Mellitus type 1 and 2* | 50 (12.7%) | 33 (13.4%) | 14 (11.2%) | 3 (13.0%) |
| *Asthma and COPD* | 30 (7.6%) | 22 (8.9%) | 8 (6.4%) | 0 |
| *Obesity* | 51 (12.9%) | 30 (12.2%) | 19 (15.2%) | 2 (8.7%) |
| *Liver disfunction* | 7 (1.8%) | 6 (2.4%) | 0 | 1 (4.3%) |
| *Kidney disfunction* | 9 (2.3%) | 9 (3.7%) | 0 | 0 |
| *Chronic hepatitis* | 6 (1.5%) | 6 (2.4%) | 0 | 0 |
| *Abdominal Aortic Aneurysm* | 4 (1.0%) | 4 (1.6%) | 0 | 0 |
| *Urolithiasis* | 9 (2.3%) | 9 (3.7%) | 0 | 0 |
| *Cholelithiasis/*<br>*Cholecystectomy* | 10 (2.5%) | 9 (3.7%) | 1 (0.8%) | 0 |
| *Polycystic Ovary Syndrome*<br>*(PCOS)* | 4 (1.0%) | 0 | 4 (3.2%) | 0 |
| *Benign Prostatic Hyperplasia*<br>*(BPH)* | 7 (1.8%) | 7 (2.8%) | 0 | 0 |
| *Irritable Bowel Syndrome* | 2 (0.5%) | 2 (0.8%) | 0 | 0 |
| *Diverticulosis* | 4 (1.0%) | 3 (1.2%) | 1 (0.8%) | 0 |
| *Fibroids* | 4 (1.0%) | 3 (1.2%) | 1 (0.8%) | 0 |
| *Ovarian cysts* | 1 (0.3%) | 0 | 1 (0.8%) | 0 |
| *Abortus provocatus* | 4 (1.0%) | 0 | 4 (3.2%) | 0 |
| *Essure device* | 1 (0.3%) | 0 | 1 (0.8%) | 0 |
| *CIN/ Loop excision procedure* | 2 (0.5%) | 0 | 2 (1.6%) | 0 |
| *Uterus extirpation* | 2 (0.5%) | 1 (0.4%) | 1 (0.8%) | 0 |
| *Vaginal prolapse* | 1 (0.3%) | 0 | 1 (0.8%) | 0 |
| *Mayer-Rokitansky-küster*<br>*syndrome* | 1 (0.3%) | 0 | 1 (0.8%) | 0 |

### Use of ultrasonography and indications

From 2019 to 2024, the number of primary care ultrasonograms performed rose from 1.1 to 9 ultrasonograms per 1000 patients. Abdominal ultrasonograms (n=246, 62.4%) were the most commonly performed, followed by gynecological (n=125, 31.7%) and other ultrasonograms (n=23, 5.8%), for instance for echo-guided interventions. The most frequent indications for abdominal ultrasonography were kidney stones and gallstones. Gynecological ultrasonograms were most often performed to assess intrauterine device (IUD) placement (35.2%).

### Findings and diagnostic accuracy

Of the 394 ultrasonograms performed, 329 (83.5%) provided conclusive results. The subsequent clinical course following the ultrasonogram is depicted in *table 2*. When evaluating the performance of primary care ultrasonograms with conclusive results, a total of 116 were found to be abnormal, warranting further investigation. Subsequent secondary care imaging indicated that one case with a serious underlying condition was missed, and 24 cases were wrongly assumed to be abnormal. In terms of overall diagnostic performance, this meant a sensitivity of 98.9% (95%CI: 94.2-100.0), specificity of 89.8% (95%CI: 85.3-93.4), positive predicting value of 79.3% (95%CI: 72.4-84.9) negative predicting value of 99.5% (95%CI: 96.8%-99.9), positive likelihood ratio of 9.73 (95%CI: 6.65-14.22) and negative likelihood ratio of 0.01 (0.00-0.08). Within the subsets of ultrasonograms performed for the two most common abdominal indications, the sensitivity for detecting gallstones was 100% (CI: 76.84-100.00%), with a specificity of 82.0% (CI: 68.56-91.42%). For kidney stones, the sensitivity was 76.9% (CI: 46.2-95.0%), and the specificity was 95.6% (CI: 87.6-99.1%).

**Table 2.** Number of ultrasonograms in four large healthcare centers in Amsterdam Southeast in 2023.

|  | *Holendrecht* | *Reigersbos* | *Klein Gooioord* | *Venserpolder* |
| --- | --- | --- | --- | --- |
| *Number of patients* | 7877 | 8329 | 7039 | 7821 |
| *Very-low income situation (%)* | 53% | 37% | 40% | 54% |
| *Number of referrals for ultrasonography in secondary care* | 104 | 189 | 166 | 197 |
| *Number of ultrasonograms in primary care* | 67 | 0 | 0 | 0 |
| *Number of secondary care ultrasonograms/ 1000 patients* | 13 | 23 | 21 | 25 |
| *Number of primary care ultrasonograms/ 1000 patients* | 9 | - | - | - |
| *Total number of ultrasonograms/ 1000 patients* | 22 | 23 | 21 | 25 |

### Impact of primary care ultrasonogram availability on overall echocardiogram usage

*Table 2* displays the number of ultrasonograms ordered by GPs per 1000 patients for four primary care centers, including the study center (Gezondheidscentrum Holendrecht) in the year 2023. The study center had a total of 22 ultrasonograms per 1000 patients ordered by GPs, of which 9 per 1000 were performed within the primary care center itself, and 13 per 1000 were performed within secondary care. The other centers had a total of 21-25 ultrasonograms per 1000 patients ordered by GPs, all of which were performed within secondary care.

## Discussion

### Key Findings

This study demonstrates the utility of ultrasonography in primary care, which displayed a gradual uptake and was used mainly to resolve cases with abdominal and gynecological complaints. The high percentage of ultrasonograms with conclusive findings, the high diagnostic accuracy particularly for ruling out clinically significant conditions, and the reduced requirement for secondary care ultrasonograms while not increasing overall GP-ordered ultrasonography consumption, suggest that ultrasonography can improve diagnostic efficiency in primary care, especially in low-income neighborhoods where access to secondary care is limited.

### Strengths and Limitations

A significant strength of this study is access to detailed data on consecutive patients in a real-world study setting. The ability to examine a large and diverse population provides robust data on the utility of ultrasonography in primary care. Limitations include the risk of verification bias as not all primary care ultrasonograms were confirmed by secondary care imaging, which may affect the diagnostic test characteristics. Additionally, while our study involves a large healthcare center, its mono-center design could affect the generalizability of our findings. Additionally, while this study illustrates the diagnostic value of primary care ultrasonography, it does not fully explore the cost-effectiveness or long-term outcomes of this practice.

### Comparison with existing literature

Studies have shown the variable uptake of ultrasonography in primary care.(1, 10) Our work concurred with these findings, with one primary care health center implementing ultrasonography into routine care while comparable practices in the vicinity did not provide such services. Our results are in line with prior studies that showed ultrasonography implementation led to substantial changes in management, fewer referrals and more frequent reassurance.(10, 11) In a recent study using point-of-care ultrasonography, the GPs’ diagnostic process and clinical decision-making was altered in nearly three out of four consultations.(11) When looking at diagnostic accuracy findings, similar outcomes have been reported compared to our study. Anderson et al performed a systematic review and found that ultrasonograms performed by GPs for kidney abnormalities had a sensitivity of 99% and specificity of 82%.(12)

### Implications for future study

Our study suggests that ultrasonographic imaging in primary care can partially replace diagnostic procedures traditionally reserved for secondary care, potentially alleviating the burden on hospitals and clinics. A promising approach to further enhance diagnostic accuracy and safety is teleconsultation, in which ultrasonographic images or short video clips, along with case descriptions, are shared with consulting radiologists. However, this requires further research to evaluate its feasibility and effectiveness. There are also concerns regarding the capacity of primary care to handle the increased diagnostic load. GPs are already under significant time pressure, and adding more diagnostic services could reduce access to other essential primary care services. Thus, the broader impact of this shift, particularly in terms of healthcare costs and operational efficiency, requires further exploration. A comprehensive cost analysis, accounting for the time and resources involved in performing ultrasonograms in primary care, would be a valuable contribution to the existing literature. This analysis should weigh the potential savings from reduced referrals against the costs of training, equipment, and the additional time required from GPs. Such an analysis was beyond the scope of the current study.

### System-Level Considerations

Healthcare actors, including policymakers, should consider the implications of expanding diagnostic services in primary care. This would involve not only assessing the immediate cost and time benefits but also determining whether the system can sustainably manage an increased workload. It is crucial that innovations do not compromise the accessibility and quality of other core healthcare services provided by GPs. To address this, integrated planning is necessary, where innovations like ultrasonogram diagnostics are introduced with the support of adequate training, resources, and adjustments in workflow to ensure primary care remains accessible and efficient for a broad range of health needs.

## Conclusion

Primary care ultrasonography offers a valuable tool for improving diagnostic decision-making within primary care and reduces the need for secondary care ultrasonography referrals. Its use in low-income neighborhoods can help bridge gaps in healthcare access. Further research is needed to explore the scalability and long-term impact of this intervention.

## Supporting information

Supplemental datafile

StaRI Checklist

## Data Availability

Data produced in the present study are available upon reasonable request to the authors

---

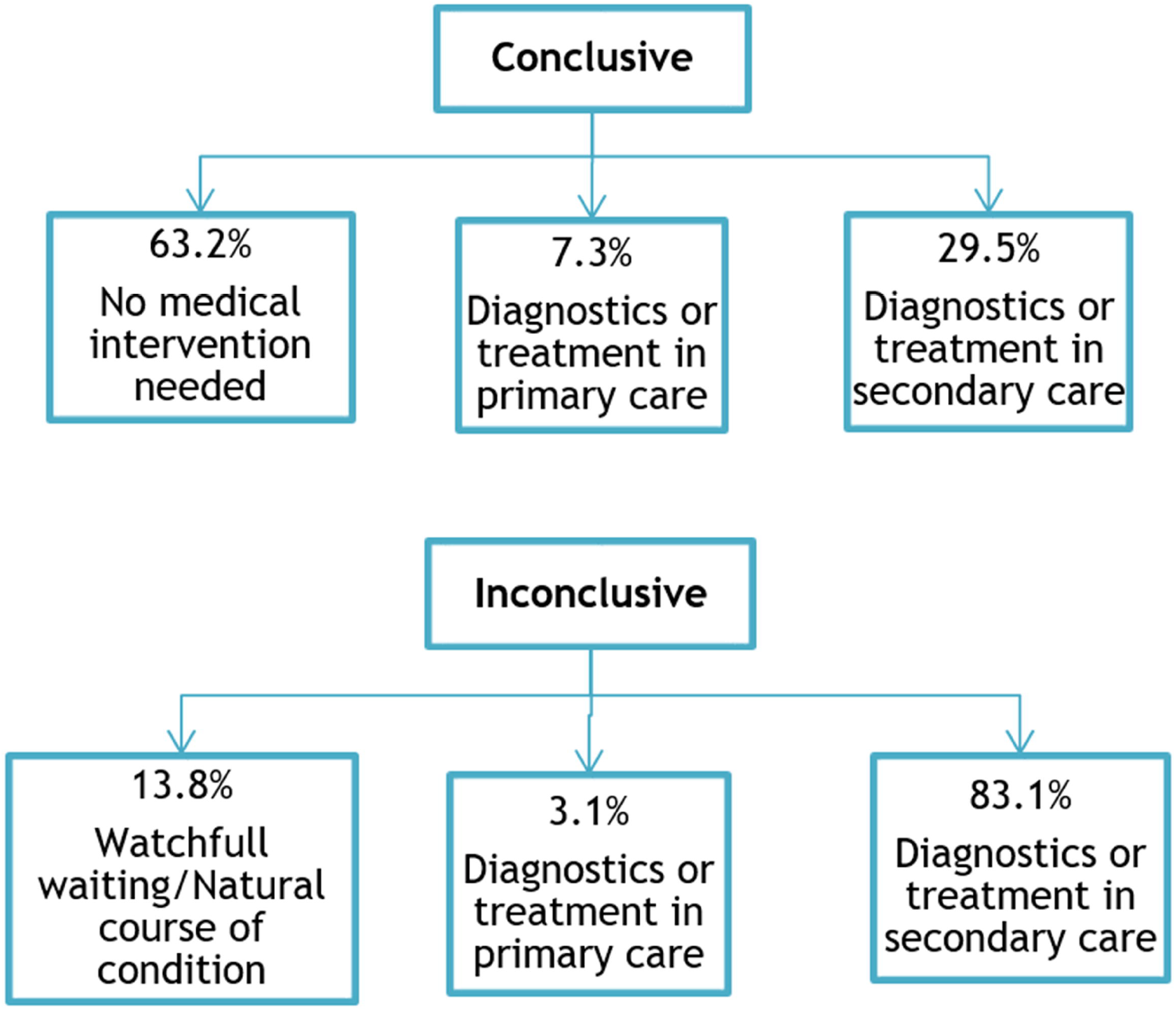

## Notes

### Competing Interest Statement

The authors have declared no competing interest.

### Funding Statement

This study did not receive any funding

### Author Declarations

The Medical Ethics Committee of Amsterdam UMC (MEC-AMC) reviewed the study and concluded that it does not fall under the scope of the Dutch Medical Research Involving Human Subjects Act (WMO). As such, the use of deidentified routine primary care data for research purposes was granted a waiver for ethical approval and informed consent (Reference: W23_074, MEC-AMC, Amsterdam).

