## Supplemental datafile for "Ultrasonography in General Practice: insights from an implementation study in a Low-income neighborhood of Amsterdam"

**Supplemental data**

| *Indications for abdominal ultrasonography* | *Frequency* | *Percentage* |
| --- | --- | --- |
| *Total* | *246* | *100%* |
| *1: Stones, obstructions, cysts or tumors of the kidneys* | 57 | 23.2% |
| *2: Stones or obstructions of the gallbladder* | 39 | 15.9% |
| *3: Tumors, metastases or other abnormalities of the liver* | 24 | 9.8% |
| *4: Bladder stones or urinary retention* | 24 | 9.8% |
| *5: Measuring aortic diameter* | 7 | 2.8% |
| *6: Fibroids* | 3 | 1.2% |
| *7: Position Intrauterine device* | 3 | 1.2% |
| *8: Other indications* | 60 | 24.4% |
| *1: Stones of the kidneys AND 2: Stones of the gallbladder* | 21 | 8.5% |
| *2: Stones of the gallbladder AND 3: Abnormalities of the liver* | 3 | 1.2% |
| *1: Stones of the kidneys AND 3: Abnormalities of the liver* | 1 | 0.4% |
| *2: Stones of the gallbladder AND 5: Measuring aortic diameter* | 1 | 0.4% |
| *1: Stones of the kidneys AND 4: Bladder stones or urinary retention* | 1 | 0.4% |
| *3: Abnormalities of the liver AND 8: Other indications* | 1 | 0.4% |
| *1: Stones of the kidneys AND 4: Bladder stones or urinary retention AND 5: Measuring aortic diameter* | 1 | 0.4% |

| *Indications for gynecological ultrasonography* | *Frequency* | *Percentage* |
| --- | --- | --- |
| *Total* | *125* | *100%* |
| *1: Position Intrauterine Device* | 44 | 35.2% |
| *2: Endometrial thickness* | 17 | 13.6% |
| *3: Fibroids* | 15 | 12.0% |
| *4: Polycystic Ovary Syndrome (PCOS)* | 11 | 8.8% |
| *5: Ovarian cysts* | 4 | 3.2% |
| *6: Other indications* | 30 | 24.0% |
| *2: Endometrial thickness AND 5: Ovarian cysts* | 2 | 1.6% |
| *2: Endometrial thickness AND 3: Fibroids* | 1 | 0.8% |
| *1: Position Intra-uterine device AND 2: Endometrial thickness AND 3: Fibroids* | 1 | 0.8% |

| *Indications for other ultrasonography* | *Frequency* | *Percentage* |
| --- | --- | --- |
| *Total* | 23 | 100% |
| *1: Subcutaneous swellings* | 17 | 74.0% |
| *2: Corpus alienum* | 3 | 13.0% |
| *3: Other indications* | 3 | 13.0% |
